# The natural history and transmission potential of asymptomatic SARS-CoV-2 infection

**DOI:** 10.1101/2020.04.27.20082347

**Authors:** Nguyen Van Vinh Chau, Vo Thanh Lam, Nguyen Thanh Dung, Lam Minh Yen, Ngo Ngoc Quang Minh, Le Manh Hung, Nghiem My Ngoc, Nguyen Tri Dung, Dinh Nguyen Huy Man, Lam Anh Nguyet, Le Thanh Hoang Nhat, Le Nguyen Truc Nhu, Nguyen Thi Han Ny, Nguyen Thi Thu Hong, Evelyne Kestelyn, Nguyen Thi Phuong Dung, Tran Chanh Xuan, Tran Tinh Hien, Nguyen Thanh Phong, Tran Nguyen Hoang Tu, Ronald B. Geskus, Tran Tan Thanh, Nguyen Thanh Truong, Nguyen Tan Binh, Tang Chi Thuong, Guy Thwaites, Le Van Tan, for OUCRU COVID-19 research group

**Affiliations:** Hospital for Tropical Diseases, Ho Chi Minh City, Vietnam; Oxford University Clinical Research Unit, Ho Chi Minh City, Vietnam; Children’s Hospital 1, Ho Chi Minh City, Vietnam; Center for Disease Control and Prevention, Ho Chi Minh City, Vietnam; Centre for Tropical Medicine and Global Health, Nuffield Department of Medicine, University of Oxford, Oxford, UK; Cu Chi Hospital, Ho Chi Minh City, Vietnam; Department of Health, Ho Chi Minh City, Vietnam

**Keywords:** COVID-19, SARS-CoV-2, coronaviruses, pandemic, Vietnam

## Abstract

**Background:** Little is known about the natural history of asymptomatic SARS-CoV-2 infection or its contribution to infection transmission.

**Methods:** We conducted a prospective study at a quarantine centre for COVID-19 in Ho Chi Minh City, Vietnam. We enrolled quarantined people with RT-PCR-confirmed SARS-CoV-2 infection, collecting clinical data, travel and contact history, and saliva at enrolment and daily nasopharyngeal throat swabs (NTS) for RT-PCR testing. We compared the natural history and transmission potential of asymptomatic and symptomatic individuals.

**Results:** Between March 10^th^ and April 4^th^, 2020, 14,000 quarantined people were tested for SARS-CoV-2; 49 were positive. Of these, 30 participated in the study: 13(43%) never had symptoms and 17(57%) were symptomatic. 17(57%) participants acquired their infection outside Vietnam. Compared with symptomatic individuals, asymptomatic people were less likely to have detectable SARS-CoV-2 in NTS samples collected at enrolment (8/13 (62%) vs. 17/17 (100%) P=0.02). SARS-CoV-2 RNA was detected in 20/27 (74%) available saliva; 7/11 (64%) in the asymptomatic and 13/16 (81%) in the symptomatic group (P=0.56). Analysis of the probability of RT-PCR positivity showed asymptomatic participants had faster viral clearance than symptomatic participants (P<0.001 for difference over first 19 days). This difference was most pronounced during the first week of follow-up. Two of the asymptomatic individuals appeared to transmit the infection to up to four contacts.

**Conclusions:** Asymptomatic SARS-CoV-2 infection is common and can be detected by analysis of saliva or NTS. NTS viral loads fall faster in asymptomatic individuals, but they appear able to transmit the virus to others.

## BACKGROUND

The rapid global spread of severe acute respiratory syndrome coronavirus-2 (SARS-CoV-2), has prompted the World Health Organization to declare a pandemic. As of April 23^rd^, 2020, more than 2.6 milllion confirmed cases and more than 180,000 deaths have been reported globally. Vietnam reported its first confirmed cases on January 22^rd^, 2020 [1]. Yet, as of April 24^th^, a total of 270 cases have been reported, with no deaths [2].

The clinical syndrome caused by SARS-CoV-2 is called COVID-19 [3], an infectious disease which varies from mild to severe, life-threatening respiratory infection. Asymptomatic infection with SARS-CoV-2 has been reported [4-6] in up to 43% of those with proven infection in a recent Italian study [7]. SARS-CoV-2 infected patients can be infectious prior to symptom (COVID-19) development and cause transmission [8, 9]. Furthermore, there is some evidence demonstrating the transmisson potential of those with RT-PCR confirmed infection who never develop symptoms during their infection (asymptomatic transmission) [4, 5, 7], suggesting asymptomatic infection may play an important role in the spread of SARS-CoV-2. SARS-CoV-2 is transmitted by respiratory droplets from infected people if they cough and/or sneeze. In the absence of respiratory symptoms, the mechanism by which asymptomatic individuals transmit SARS-CoV-2 to their contacts remains unclear. In most countries, only patients with moderate or severe disease are admitted to hospital for management [10-13], leaving those without symptoms, or with mild disease, uncharacterized, especially concerning their laboratory and virological findings.

We therefore studied asymptomatic individuals with SARS-CoV-2 infection and those with mild disease identified as part of ongoing contact-tracing and airport quarantine implemented in Ho Chi Minh City (HCMC), Vietnam. Our aims were to compare the duration of viral detection and abundance in the respiratory tract, including saliva, of asymptomatic and mildly symptomatic patients, and assess their ability to transmit the virus to others.

## MATERIALS AND METHODS

### Vietnam contaiment approach

Since January 2020, various control measures, including isolation of confirmed cases, contact-tracing, airport quarantine and social distancing have been implemented in Vietnam with increasing stringency as the pandemic progressed worldwide (**Figure 1**) [14, 15]. Accordingly, anyone known to have been in contact with a confirmed COVID-19 case, or having travelled to Vietnam from a COVID-19 affected country, were isolated for ≥14 days at a designated isolation center.

**Figure 1:**
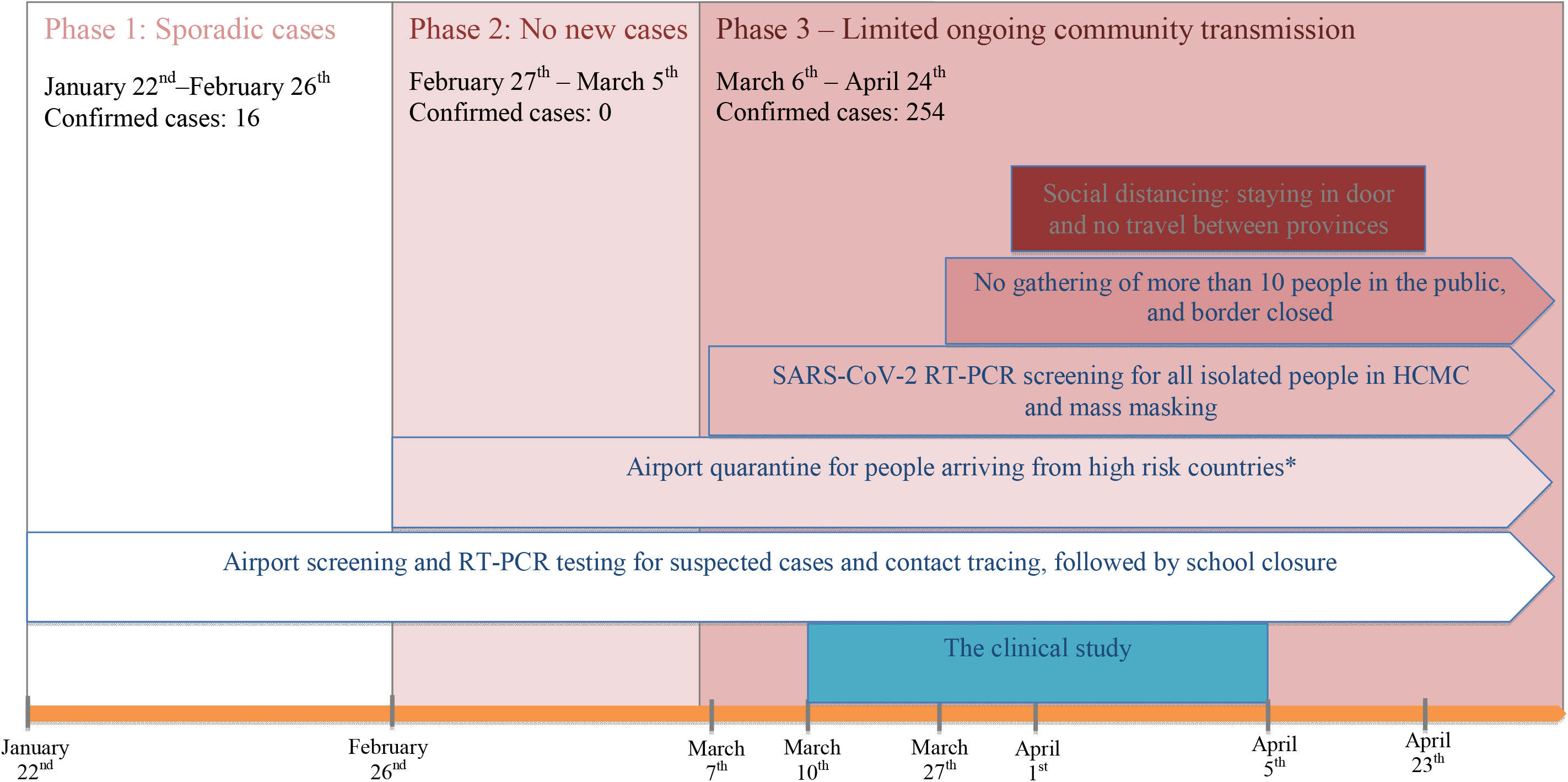
Timelines of containment strategies applied in Vietnam and in Ho Chi Minh City since the beginning of 2020 as the epidemic/pandemic progresses alongside the implementation of SARS-CoV-2 RT-PCR testing and the duration of the clinical study. **Note to figure 1**: *Initially China, followed by Korea, other European counties (Italy, France, UK etc) and the USA

From the second week of March 2020, all isolated individuals were subject to serial SARS-CoV-2 nasopharyngeal throat swab (NTS) screening by real time RT-PCR. A confirmed case was established if two independent RT-PCR assays (E gene and RdRP RT-PCR assays) were positive [16]. Confirmed cases were admitted to a designated COVID-19 hospital for follow-up until they recovered and/or had at least two consecutive days with negative SARS-CoV-2 RT-PCR NTS [17].

### Setting

The Hospital for Tropical Diseases (HTD) is a tertiary referral infectious diseases hospital responsible for receiving and treating patients with COVID-19 in southern Vietnam. From January 2020 to the first week of April, HTD was responsible for RT-PCR screening of 80% of quarantined people in HCMC.

In addition to its main campus in the centre of HCMC, HTD has two designated 300-bed centres for the care of confirmed/suspected cases with COVID-19, namely Cu Chi and Can Gio Hospitals, located approximately 60 km to the West and East, respectively, of HCMC (**Figure 2A**). The present study was conducted at Cu Chi Hospital.

**Figure 2:**
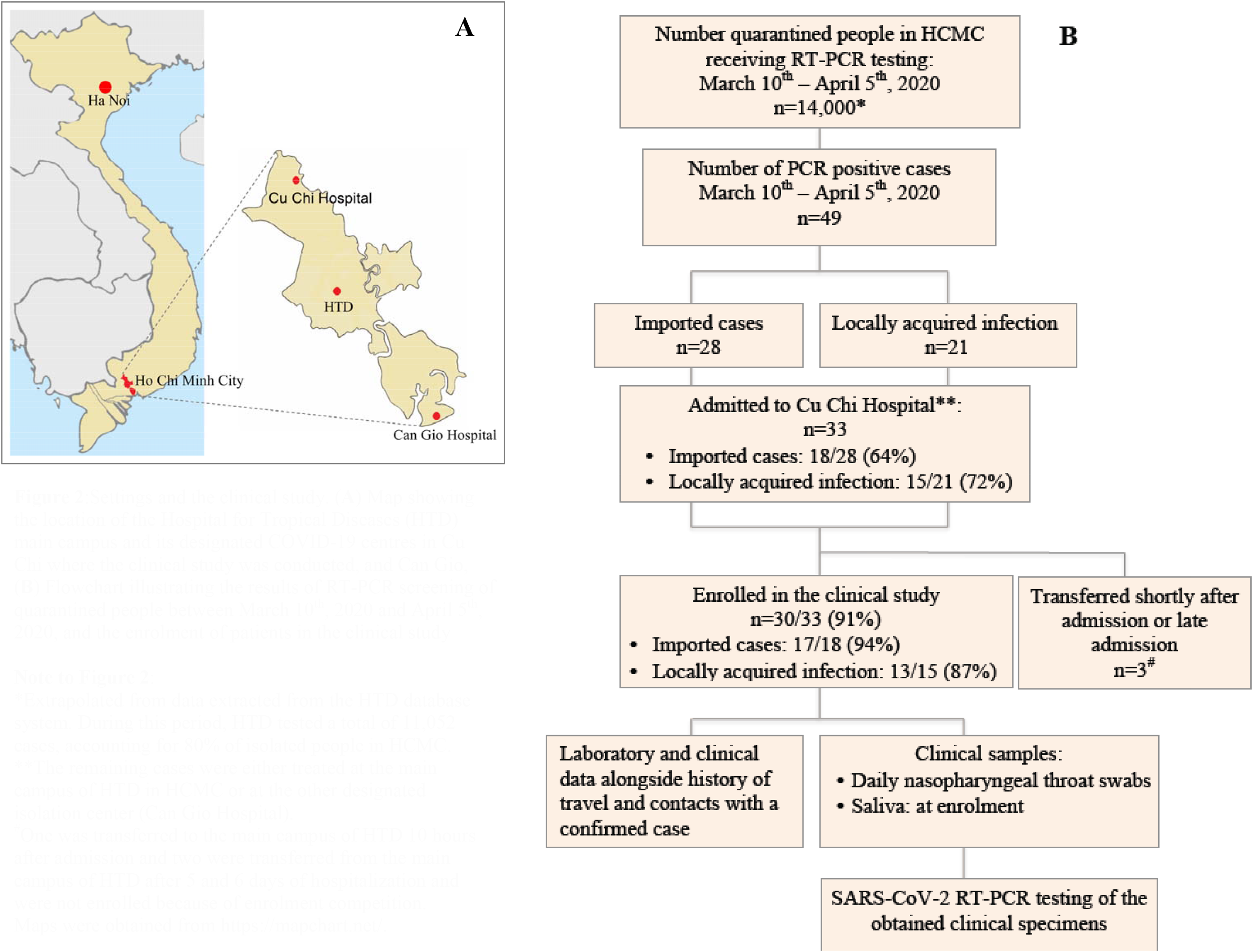
Settings and the clinical study.(**A**) Map showing the location of the Hospital for Tropical Diseases (HTD) main campus and its designated COVID-19 centres in Cu Chi where the clinical study was conducted, and Can Gio, Flowchart illustrating the results of RT-PCR screening of quarantined people between March 10^th^, 2020 and April 5^th^, 2020, and the enrolment of patients in the clinical study **Note to Figure 2**: *Extrapolated from data extracted from the HTD database system. During this period, HTD tested a total of 11,052 cases, accounting for 80% of isolated people in HCMC. **The remaining cases were either treated at the main campus of HTD in HCMC or at the other designated isolation center (Can Gio Hospital). ^#^One was transferred to the main campus of HTD 10 hours after admission and two were transferred from the main campus of HTD after 5 and 6 days of hospitalization and were not enrolled because of enrolment competition. Maps were obtained from https://mapchart.net/.

### Patient erollment and data collection

We enrolled individuals with confirmed SARS-CoV-2 infection admitted to Cu Chi Hospital from March 10^th^ to April 4^th^, 2020. From each participant, we prospectively collected demographic and clinical data, travel history and information concerning contact with confirmed COVID-19 cases, using standardized paper case record forms.

We collected NTS, combining them into a single tube containing 1ml of viral transport medium. NTS were taken daily from enrolment to hospital discharge (**Figure 2B**). Additionally, a saliva sample was obtained at enrolment. After collections, clinical samples were stored at −4°C at the study site and were then transferred to the HTD laboratory in HCMC within 4 hours for analysis.

### Viral RNA extraction and SARS-CoV-2 RT-PCR analysis

We manually extracted viral RNA from 140 ul of NTS and saliva samples (if volume was sufficient for testing) using the QIAamp viral RNA kit (QIAgen GmbH, Hilden, Germany), and then recovered the cleaned up RNA in 50 ul of elution buffer provided with the kit. Since we enroled patients who had a confirmed diagnosis by two independent RT-PCRs (Egene and RdRp assays) as per the WHO recomendation [16], we used E gene assay for testing of samples collected from enrolment onward. Real time RT-PCR were carried out as previously described [16].

### Data analysis

For viral-load associated analysis, in the absence of quantitative RT-PCR results, we use cycle threshold (Ct) values as surrogates. We used the t-test to compare the difference in measured Ct values obtained at enrolment between the two groups, Wilcoxon signed-rank test to compare the measured Ct values between NST and saliva, and chi-square test to compare two proportions. We compared the trend in the detection probability of SARS-CoV-2 and the viral RNA load in NTS between asymptomatic and symptomatic individuals. For the detection probability, we fitted a logistic regression model that quantifies the probability to test positive over time. We used generalized estimating equations (geepack package in R [18]) to correct for the repeated measurements per individual. We assumed that those who left the study earlier, after several days with a negative test result, remained negative until day 19. For the trend in Ct, we used a zero-inflated mixed effects model for semi-continuous data (GLMMadaptive package in R). Since the Ct data have an upper threshold of 40.5, we used the transformation Y=40.5-Ct. Hence, an undetectable viral load was given the value zero. We report the mean value on the original scale, which is a weighted combination of the value 40.5 for those that test negative and the measured value of those that test positive. Additionally, we compared the measured Ct values between the two groups; for this we used a random effects model. Further details can be found in the supplementary code. Note that inference with respect to measured Ct values should be interpreted with caution, because the subgroup of negative values is selectively excluded. Apart from R, we also performed the analysis in SPSS V23.0 (IBM Corp, NY, US) and generated the figures using GraphPad PRISM^®^ V5.04 (GraphPad Software Inc, CA, US) and R.

### Ethics

This clinical study received approvals from the Institutional Review Board of the HTD and the Oxford Tropical Research Ethics Committee of the University of Oxford. Study participants gave their written informed consent.

## RESULTS

### RT-PCR screening of quarantined people

Between March 10^th^ and April 4^th^, 2020, approximately 14,000 people were referred to one of nine designated quarantine centres deployed across HCMC, and were screened for SARS-CoV- 2 by RT-PCR of NTS. Forty-nine people had a positive test, accounting for 96% (49/51) of all reported cases in HCMC during the same period. The other two self-presented to local hospitals after falling ill. Of these 49, 33 (67%) were admitted to Cu Chi Hospital, and 30 (30/33, 91%) agreed to participate in the clinical study (**Figure 2B**).

### Baseline characteristics of the study participants

Of the 30 study participants, 16 were imported cases (i.e. they acquired the infection outside of Vietnam, **Supplementary Figure 2**), and 14 acquired the infection locally; all 14 had an epidemiological link with two community transmission clusters occurring in HCMC during the study period. Of the locally acquired infections, 7 (50%) were asymptomatic; while 6 (38%) of the imported cases were asymptomatic. Those with locally acquired infection were more likely to be male than those with infection acquired outside of Vietnam (**Table 2**).

Seventeen of the participants had mild respiratory disease; i.e. no requirement for supplementary oxygen during hospitalization. None of the participants developed severe disease. The other 13 patients had no symptoms or signs of infection throughout their hospital admission. The demographic and laboratory characteristics of the two groups were similar at enrolment (**Table 1 and 2**). A small proportion of symptomatic patients presented with diarrhea and/or lost their sense of smell. None of the 30 participants had abnormal findings on chest radiographs.

**Table 1:**
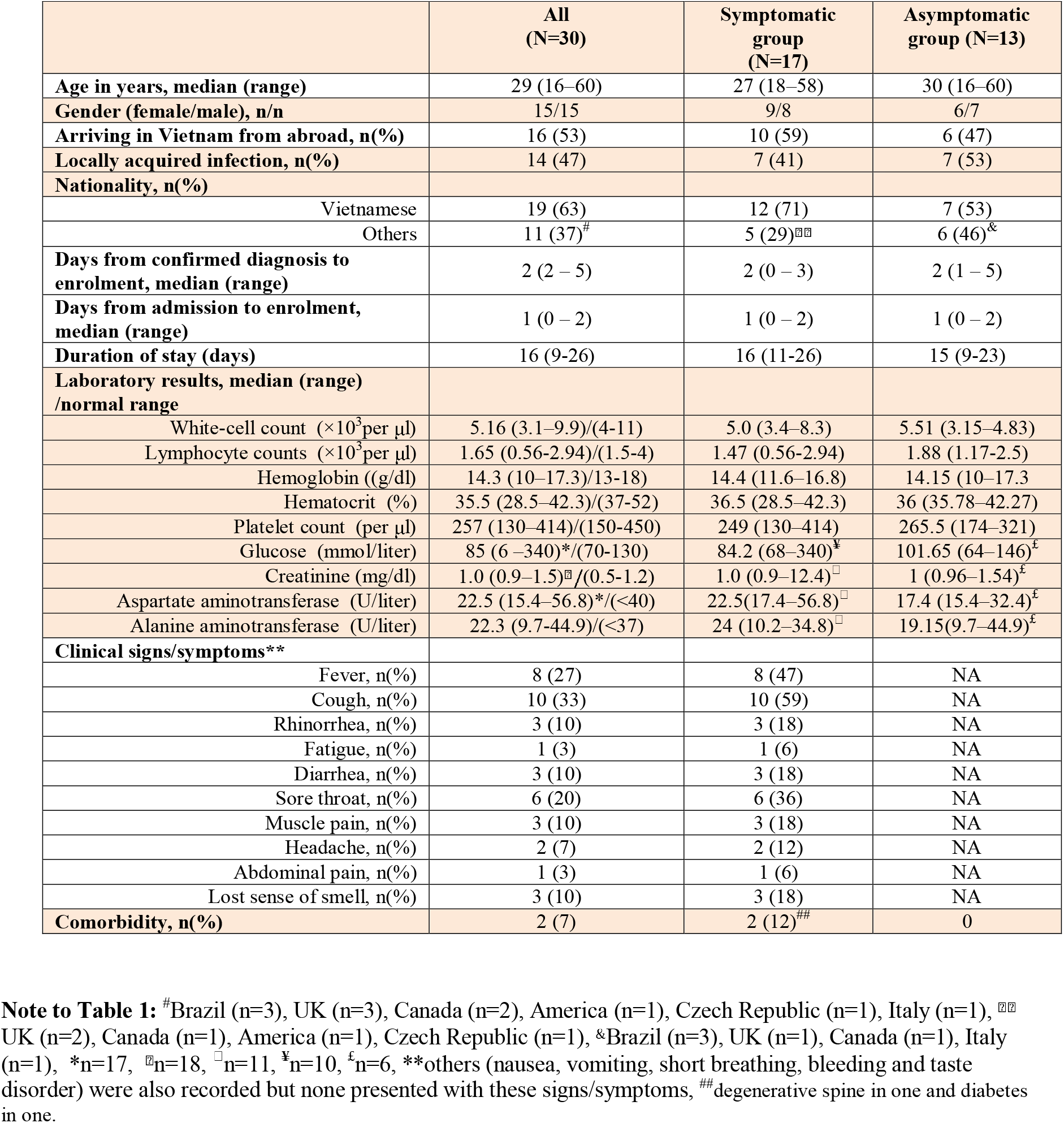
Baseline characteristics of the study participants

**Table 2:**
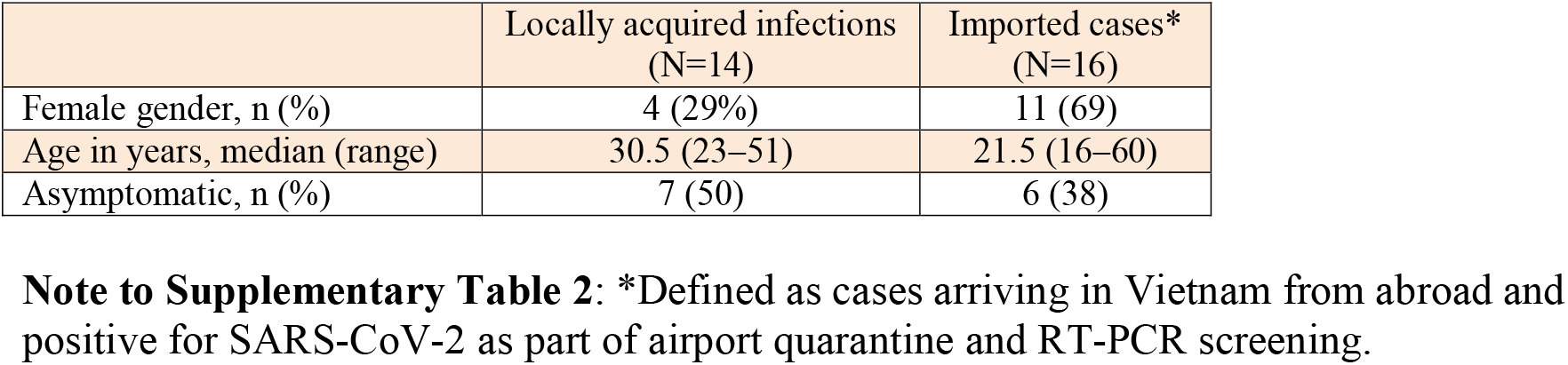
Demographic of imported cases and cases of locally acquired infection

|  | Locally acquired infections<br>(N=14) | Imported cases*<br>(N=16) |
| --- | --- | --- |
| Female gender, n (%) | 4 (29%) | 11 (69) |
| Age in years, median (range) | 30.5 (23–51) | 21.5 (16–60) |
| Asymptomatic, n (%) | 7 (50) | 6 (38) |

**Note to Supplementary Table 2:** \*Defined as cases arriving in Vietnam from abroad and positive for SARS-CoV-2 as part of airport quarantine and RT-PCR screening.

### Viral detection in NTS and saliva samples

Compared with symptomatic patients, those with asymptomatic infection were less likely to have detectable SARS-CoV-2 in NTS samples collected at enrolment (8/13 (62%) vs. 17/17 (100%) P=0.02). However, 4/5 patients whose NTS collected at enrolment were negative had an NTS positive result in one of the subsequent sampling days, but with a high Ct value (**Supplementary Figure 2**), suggesting these patients had low viral load in their respiratory samples.

Of the 30 study participants, 27 (90%) had a saliva sample collected at enrollment with sufficient volume for RT-PCR analysis. SARS-CoV-2 RNA was detected in 20/27 (74%) available saliva, 7/11 (64%) in the asymptomatic and 13/16 (81%) in the symptomatic group (P=0.56). There was one patient, whose NTS collected at enrolment was negative but saliva was positive. Accordingly, a combination of both NTS and saliva samples collected at enrolment slightly increased the diagnostic yield of samples collected at enrolment of the asymptomatic group

### Quantification of viral RNA in NTS and saliva at enrolment

At enrolment, among those who were RT-PCR positive, the viral loads measured in NTS and saliva were similar in asymptomatic and symptomatic patients (**Figure 3A**). However, among asymptomatic patients who had both saliva and NTS collected, higher viral load was observed in NTS than in saliva (P=0.031) (**Figure 3B**). A similar trend was observed for symptomatic cases (P=0.064) (**Figure 3B**).

**Figure 3:**
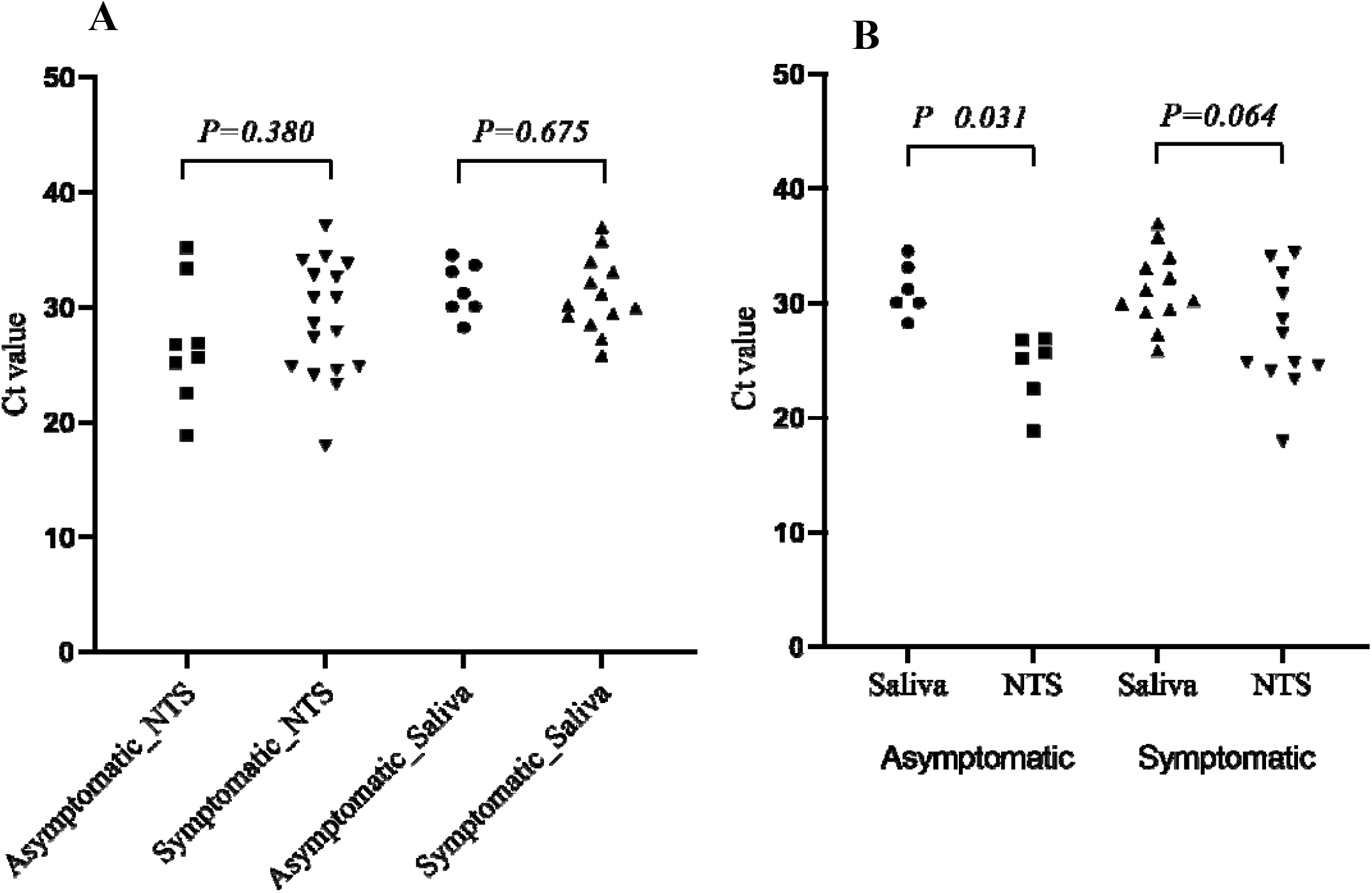
Comparison of Ct values of SARS-CoV-2 real time RT-PCR obtained from nasopharyngeal swabs (NTS) and saliva. (**A**) Data include results of RT-PCR analysis of all available NTS and saliva collected from 30 participants at enrolment, (**B**) Data only include results of RT-PCR analysis of paired NTS and saliva of 6 asymptomatic and 12 symptomatic patients who had both sample types collected at enrolment.

### Quantification of viral RNA and duration of viral detection in NTS

During follow-up, Ct-values differed between the two groups (P=0.027 for difference over first 19 days, **Figure 4A**), with asymptomatic patients having lower viral load than the symptomatic patients. If restricted to RT-PCR positive samples, viral RNA abundance was similar to slightly lower in the asymptomatic participants (P=0.086) (**Supplementary Figure 3**).

Analysis of the probability of RT-PCR positivity showed asymptomatic participants had a lower probability of having a positive RT-PCR result (i.e. a faster viral clearance) than symptomatic participants (P<0.001 for difference over first 19 days, **Figure 4B**). This difference was most pronounced during the first week of follow-up. After this period, the probability of detection quickly fell to almost zero in both groups. The majority of the positive patients were weakly positive (Ct value >32) in this period.

**Figure 4:**
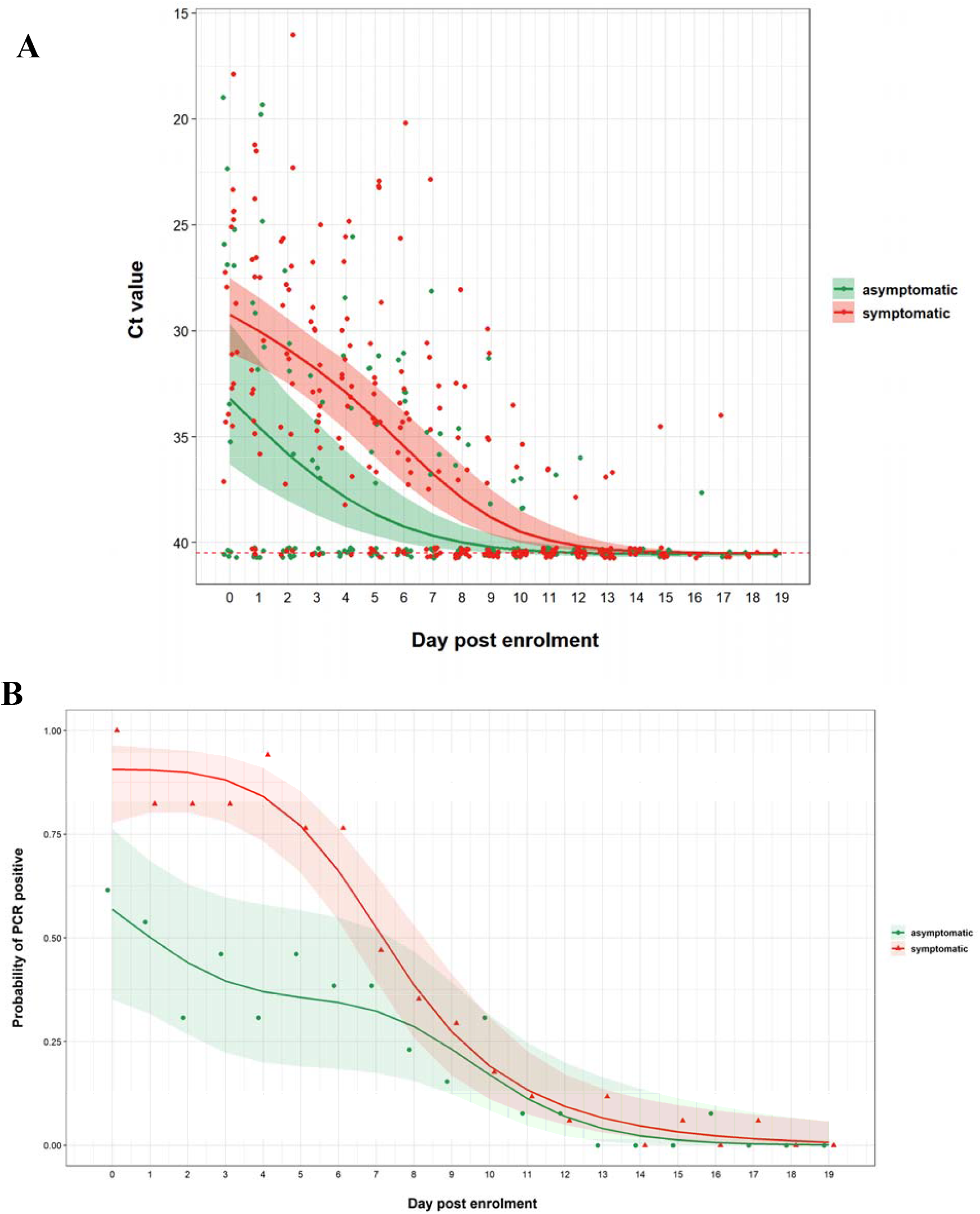
Trends in Ct values and viral detection probability in nasopharyngeal throat swabs over the course of hospitalization; (**A**) Changes of Ct values, relatively reflect the level of viral load, (B) Dynamics of viral detection probability from enrolment onward **Note to Figure 4**: Each dot represents one observed value (A) or the mean value (i.e. frequency) per day (B), lines indicate mean and shades indicate 95% confidence interval.

### Presumed transmission from asymptomatic carriers

Fourteen participants were identified to have an epidemiological link with two community-transmission clusters occurring in HCMC during the study period. Cluster #1 had three patients participating in the present study. Of these three, 2 had contact with a confirmed case on March 2^nd^, who was not enrolled in this study because this patient was admitted to a different hospital. Subsequently, one developed fever, runny nose and sore throat on March 12^th^, 2020, suggesting an incubation period of 10 days, and tested positive for SARS-CoV-2 on March 13^rd^, 2020. The other had no fever or any signs/symptoms suggestive of infection and was positive for SARS-CoV-2 on March 14, 2020. Two days later, a colleague of these two cases developed mild respiratory symptoms, including runny nose and loss of sense of smell, and tested positive for SARS-CoV-2 on March 17^th^, 2020.

Cluster #2 included 11 study participants, including 7 with asymptomatic infection (**Figure 5**). We identified a transmission chain involving an asymptomatic participant (patient #19) who was positive for SARS-CoV-2 on March 25 (Ct value of NTS: 24, and saliva: 28). Subsequently, a contact of this case (patient #22) was positive for SARS-CoV-2 on March 23^rd^ (Ct value of NTS: 23, and saliva: 34), although this contact did not develop symptoms. Furthermore, on March 27^th^, a contact of both patient #19 and #22 (patient #27) presented with cough and sore throat, with positive NTS for SARS-COV-2. Additionally, patient #26, contact of patient #22, who was also a contact of patient #19, was confirmed with SARS-CoV-2 on March 20^th^, also without any symptoms. An additional transmission chain from cluster #2 was recorded between patient #24 and #29, both of whom were asymptomatic (**Figure 5**).

**Figure 5:**
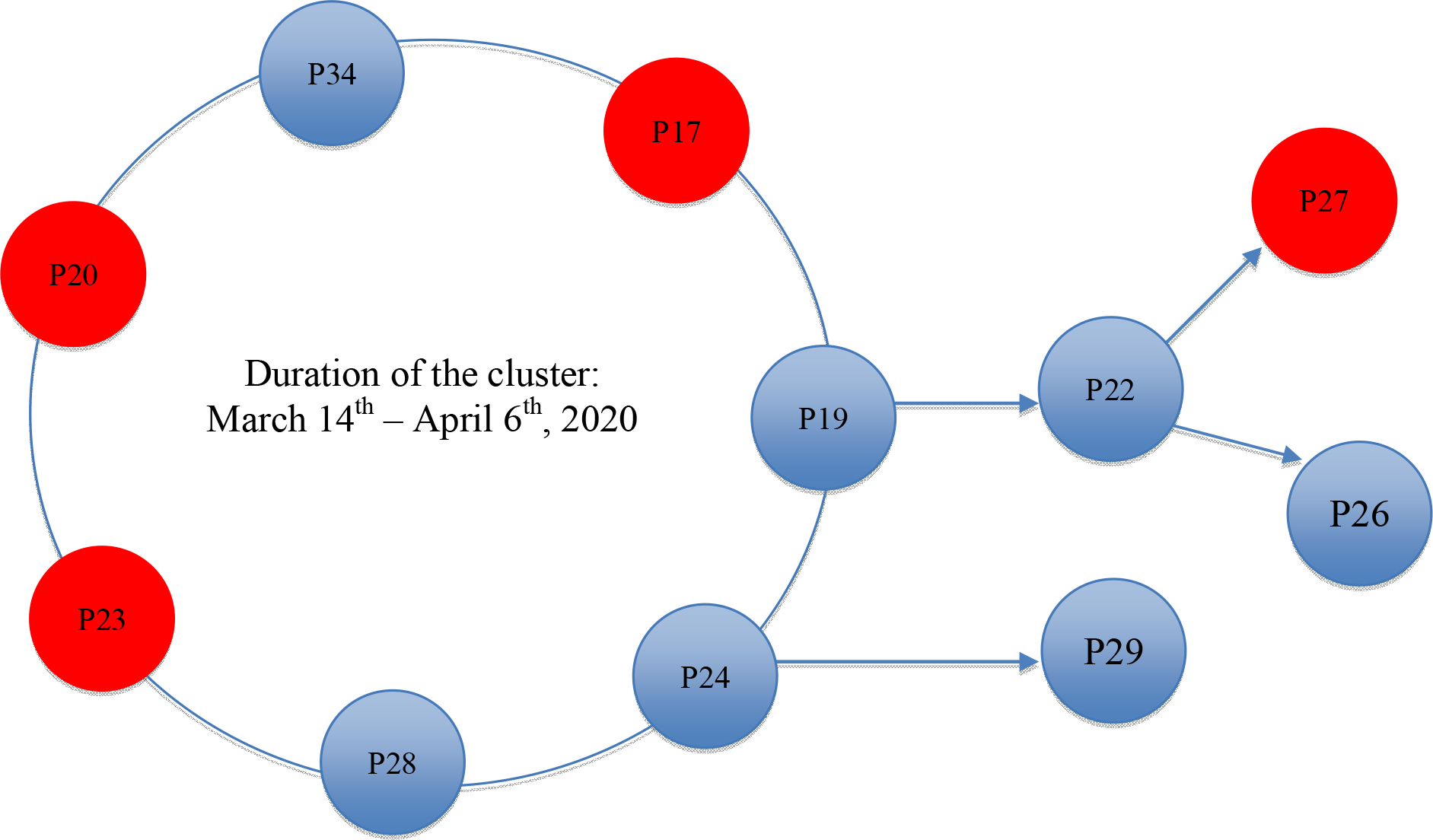
Illustration of cases with an epidemiological link with community transmission cluster #2. Red circles indicate symptomatic patients, and blue circles indicate asymptomatic cases. Patients sitting on the open circle are people of the cluster who first had contacts with each other on the same day. Patient numbers correspond to the numbers presented in Supplementary Figure 2.

## DISCUSSION

Despite the rapid global spread of SARS-CoV-2, community transmission of SARS-CoV-2 in Vietnam remains exceptionally low [2]. Indeed, while the first reported cases date back to January 23^rd^, 2020, as of April 24^th^, there have been only 270 reported cases in Vietnam, including 170 imported cases and 100 cases acquired locally [2, 19]. During the same period, the number of confirmed cases worldwide increased from 582 to more than 2.7 million.

Social distancing, school closure, isolation of confirmed cases and their contacts, and airport quarantine [14, 15] coupled with RT-PCR testing for all the isolated people have been the main measures leading to the current success of Vietnam’s COVID-19 control [14, 15]. The quarantine of large numbers of contacts has offered a unique opportunity to study the natural history of SARS-CoV-2 infection, especially in those without symptoms.

Using data from 30 patients, representing 56% of the reported case in HCMC since the beginning of the epidemic, we provide important insights into the natural history of SARS-CoV-2 infection. We found that 43% of SARS-CoV-2 positive cases were asymptomatic, with comparable detection rates and viral load of SARS-CoV-2 in saliva between symptomatic and asymptomatic cases. However, at enrolment and during follow up asymptomatic individuals had a lower probablity of having a postive RT-PCR diagnosis and lower viral load in NTS. Yet, despite these data suggesting faster viral clearance from the respiratory tract, we found good evidence these asymptomatic individuals transmitted the virus to others.

SARS-CoV-2 RNA has previously been detected in saliva of COVID-19 patients [20, 21], demonstrating the utility potential of easy-to-collect saliva samples for the diagnosis of COVID-19 [22]. However, to the best of our knowledge, detection of SARS-CoV-2 in saliva of asymptomatic cases has not been reported. Slightly higher viral loads (lower Ct values in **Figure 3**) were found in NTS than saliva, but saliva is an easier specimen to collect and may represent a better sample for mass disease-screening programmes. The ease of detecting virus in the saliva is also consistent with the known high infectiousness of SARS-CoV-2 and its ready ability to spread through droplet transmission even without respiratory symptoms.

Although the viral loads at enrolment were similar between the asymptomatic and symptomatic participants (if restricted to the positive cases), the virus appeared to be cleared faster from the respiratory tract in asymptomatic people. These differences suggest symptoms and subsequent disease severity may depend on the size of the infectious viral inoculum and/or an individual’s ability to clear the infection. However, we cannot also rule out that the time from infection to sample collection was longer in asymptomatic individuals. Other reasons for asymptomatic infection include pre-existing cross-immunity as a consequence of previous exposure to common human coronavirus, which may enhance immunity and control of the infection in some individuals [23].

Nevertheless, despite faster viral clearance in asymptomatic individuals, we found good evidence that they were still able to transmit the infection. Two of the asymptomatic participants were the highly likely origin of at least 2, and possible 4 further infections. Transmission from asymptomatic and especially pre-symptomatic individuals has been suggested previously [4-6, 8, 9] and may explain why the virus is so hard to control. The finding supports the Vietnam approach of vigorous case-finding, quarantining, and testing and suggests they are essential of the infection is to be controlled.

The strengths of our study include the inclusion of the majority of asymptomatic and symptomatic cases reported in southern Vietnam over 4 weeks, without selection bias based on symptoms or disease severity. In so doing, we were able to study prospectively the full spectrum of SARS-CoV-2 infection. Our study also has some limitations. We did not perform virus culture to demonstrate the infectiousness of SARS-CoV-2 detected by RT-PCR in saliva, although through contact history, we identified at least two transmission events from completely asymptomatic individuals. Additionally, we did not perform chest computerized tomography scans [24], which are more sensitive than chest radiographs for the detection of lung abnormalities. Therefore, we may have underestimated the sub-clinical findings of SARS-CoV-2 infection. Lastly, none of the participants developed severe disease. However, as of April 23, 2020, only three severe COVID-19 cases have been reported in HCMC and there have, as yet, been no COVID-19 related deaths in Vietnam.

To summarize, we demonstrate that a high proportion (43%) of quarantined people who were RT-PCR positive for SARS-CoV-2 were asymptomatic. These individuals carried SARS-CoV- 2 in their respiratory tract and saliva, and were potentially contagious. They would not have been identified without the control measures as currently applied in Vietnam. Therefore, our findings emphasize the importance of contact tracing, airport quarantine and RT-PCR screening for SARS-CoV-2 among isolated people in controlling the ongoing pandemic.

## Data Availability

R Codes used and additional information were provided in supplementary materials.

## ACKNOWLEDGEMENTS

This study was funded by the Wellcome Trust of Great Britain (106680/B/14/Z and 204904/Z/16/Z).

We are indebt to Dr Vu Thi Ty Hang, Dr Phan Nguyen Quoc Khanh, Ms Nguyen Thanh Ngoc, Ms Le Kim Thanh, and the OUCRU IT/CTU departments, especially Mr Ho Van Hien and Dr Nguyen Than Ha Quyen, for their support.

We thank Professor Lin-Fa Wang and Dr Danielle Anderson of Duke-NUS Medical School, Singapore for their guidance in the development of diagnostic reagents in this study.

We thank the patients for their participations in this study and the doctors and nurses of Cu Chi Hospital, who cared for the patients and provided the logistic support with the study.

## OUCRU COVID-19 Research Group

### Hospital for Tropical Diseases, Ho Chi Minh City, Vietnam

Nguyen Van Vinh Chau, Nguyen Thanh Dung, Le Manh Hung, Huynh Thi Loan, Nguyen Thanh Truong, Nguyen Thanh Phong, Dinh Nguyen Huy Man, Nguyen Van Hao, Duong Bich Thuy, Nghiem My Ngoc, Nguyen Phu Huong Lan, Pham Thi Ngoc Thoa, Tran Nguyen Phuong Thao, Tran Thi Lan Phuong, Le Thi Tam Uyen, Tran Thi Thanh Tam, Bui Thi Ton That, Huynh Kim Nhung, Ngo Tan Tai, Tran Nguyen Hoang Tu, Vo Trong Vuong, Dinh Thi Bich Ty, Le Thi Dung, Thai Lam Uyen, Nguyen Thi My Tien, Ho Thi Thu Thao, Nguyen Ngoc Thao, Huynh Ngoc Thien Vuong, Pham Ngoc Phuong Thao, Phan Minh Phuong

### Oxford University Clinical Research Unit, Ho Chi Minh City, Vietnam

Dong Thi Hoai Tam, Evelyne Kestelyn, Donovan Joseph, Ronald Geskus, Guy Thwaites, H. Rogier van Doorn, Huynh Le Anh Huy, Huynh Ngan Ha, Huynh Xuan Yen, Jennifer Van Nuil, Jeremy Day, Joseph Donovan, Katrina Lawson, Lam Anh Nguyet, Lam Minh Yen, Le Nguyen Truc Nhu, Le Thanh Hoang Nhat, Le Van Tan, Sonia Lewycka Odette, Louise Thwaites, Maia Rabaa, Marc Choisy, Mary Chambers, Motiur Rahman, Ngo Thi Hoa, Nguyen Thanh Thuy Nhien, Nguyen Thi Han Ny, Nguyen Thi Kim Tuyen, Nguyen Thi Phuong Dung, Nguyen Thi Thu Hong, Nguyen Xuan Truong, Phan Nguyen Quoc Khanh, Phung Le Kim Yen, Sophie Yacoub, Thomas Kesteman, Nguyen Thuy Thuong Thuong, Tran Tan Thanh, Tran Tinh Hien, Vu Thi Ty Hang

### Center for Disease Control and Prevention, Ho Chi Minh City, Vietnam

Nguyen Tri Dung, Le Hong Nga

## SUPPLEMENTARY MATERIALS

**Supplementary Figure 1:**
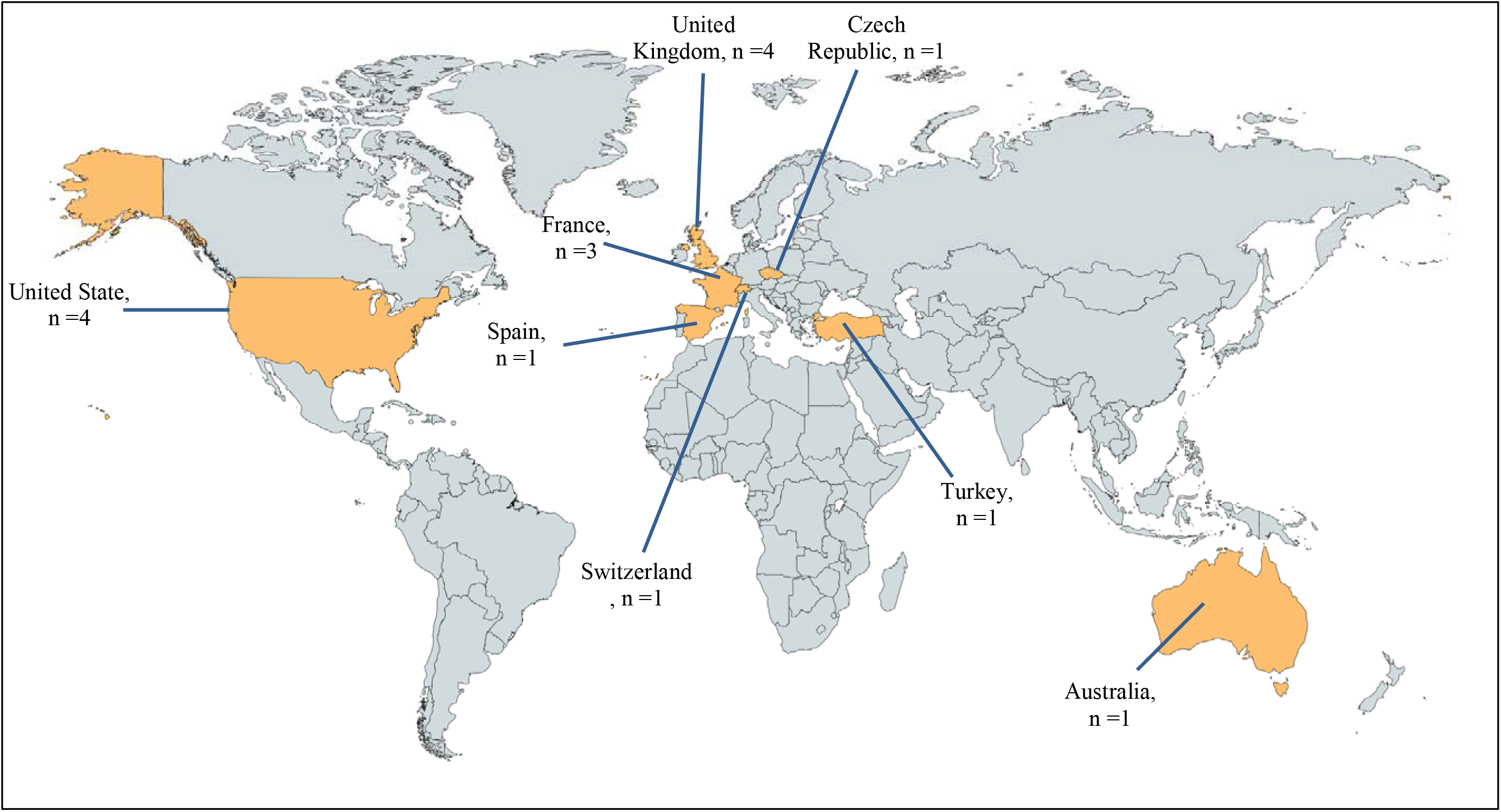
Map showing countries where the imported cases stayed before travelling to Vietnam **Note to supplementary Figure 1**: Maps were obtained from https://mapchart.net/

**Supplementary Figure 2:**
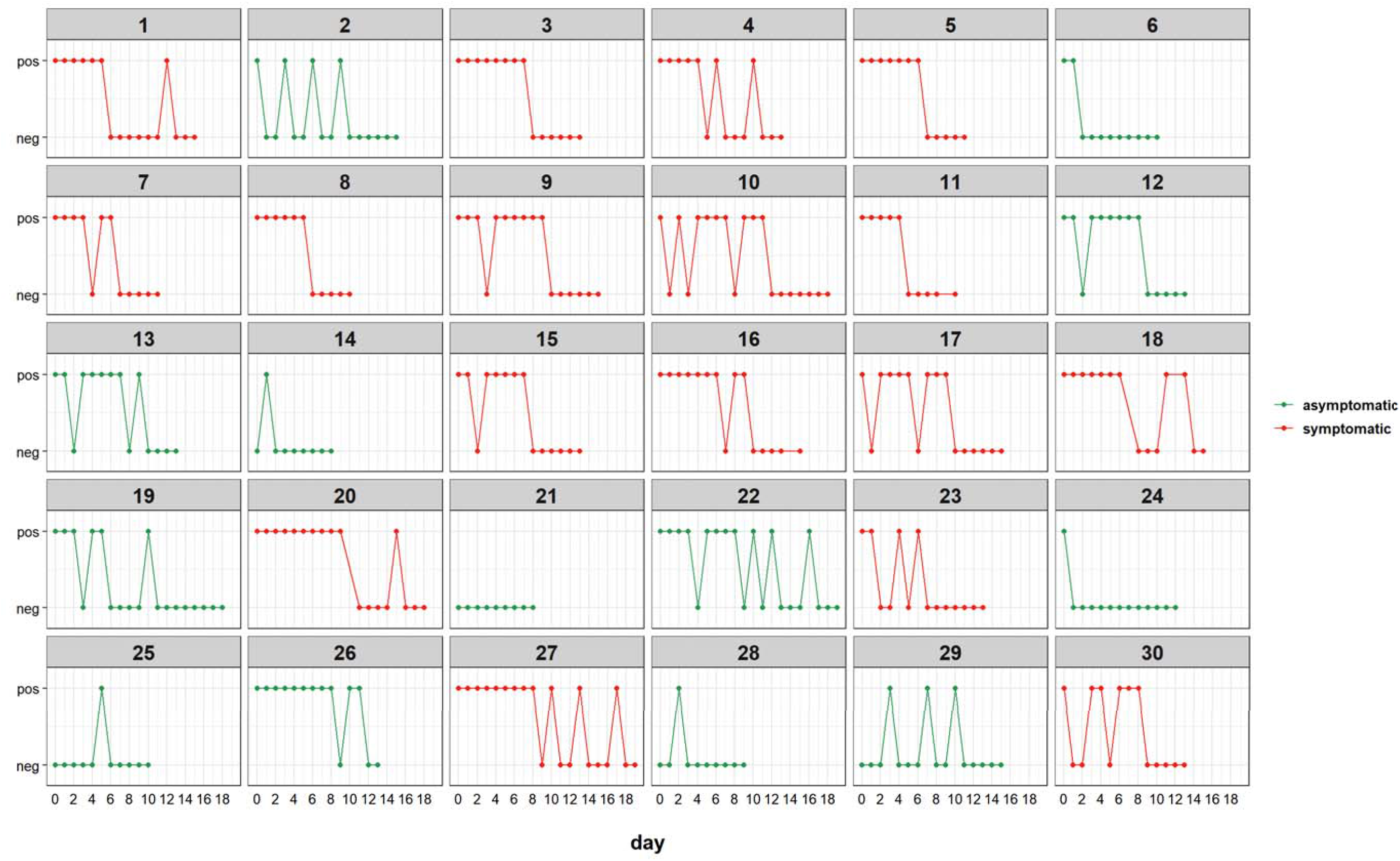
Individuals data on Ct values of SARS-CoV-2 real-time RT-PCR obtained from analysis of nasopharyngeal throat swabs collected at enrollment and during follow-up **Note to Supplementary Figure 2**: Horizontal dash lines indicate the assay cut off.

**Supplementary Figure 3:**
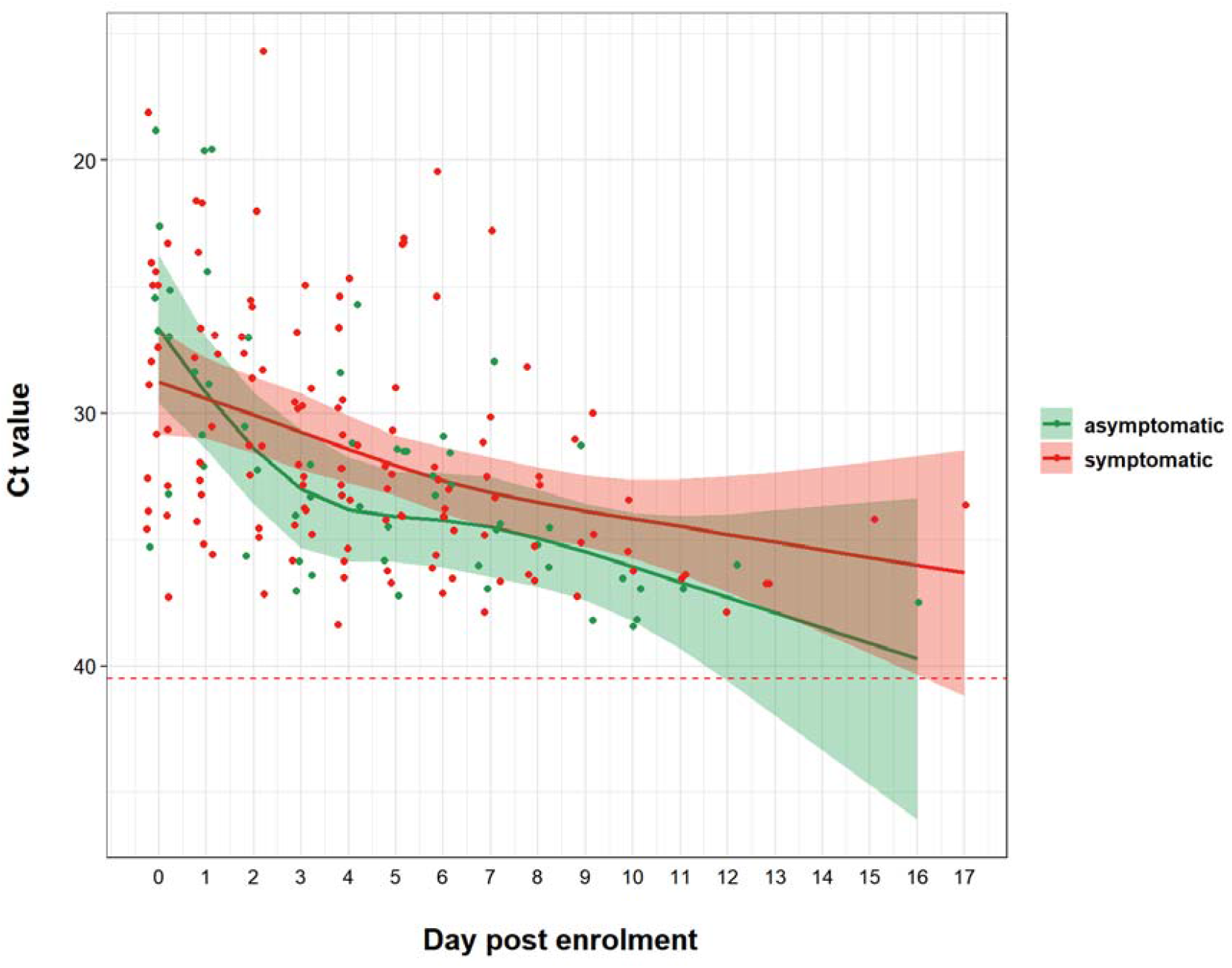
Dynamics of Ct values during the course of quarantine or hospitalization in nasopharyngeal throat swabs of people who remained RT-PCR positive.

## Notes

### Competing Interest Statement

The authors have declared no competing interest.

### Funding Statement

Wellcome Trust grants: 106680/B/14/Z and 204904/Z/16/Z

## REFERENCES

1. Phan, L.T., T.V. Nguyen, Q.C. Luong, T.V. Nguyen, H.T. Nguyen, H.Q. Le, T.T. Nguyen, T.M. Cao, and Q.D. Pham, Importation and Human-to-Human Transmission of a Novel Coronavirus in Vietnam. N Engl J Med, 2020. 382(9): p. 872–874.

2. Ministry of Health, Vietnam. https://ncov.moh.gov.vn/.

3. Zhu, N., D. Zhang, W. Wang, X. Li, B. Yang, J. Song, X. Zhao, B. Huang, W. Shi, R. Lu, et al., A Novel Coronavirus from Patients with Pneumonia in China, 2019. N Engl J Med, 2020. 382(8): p. 727–733.

4. Bai, Y., L. Yao, T. Wei, F. Tian, D.Y. Jin, L. Chen, and M. Wang, Presumed Asymptomatic Carrier Transmission of COVID-19. JAMA, 2020.

5. Chen, L., Q. Li, D. Zheng, H. Jiang, Y. Wei, L. Zou, L. Feng, G. Xiong, G. Sun, H. Wang, et al., Clinical Characteristics of Pregnant Women with Covid-19 in Wuhan, China. N Engl J Med, 2020.

6. Gudbjartsson, D.F., A. Helgason, H. Jonsson, O.T. Magnusson, P. Melsted, G.L. Norddahl, J. Saemundsdottir, A. Sigurdsson, P. Sulem, A.B. Agustsdottir, et al., Spread of SARS-CoV-2 in the Icelandic Population. N Engl J Med, 2020.

7. Lavezzo, E., E. Franchin, C. Ciavarella, G. Cuomo-Dannenburg, L. Barzon, C. Del Vecchio, L. Rossi, R. Manganelli, A. Loregian, N. Navarin, et al., 2020.

8. Wei, W.E., Z. Li, C.J. Chiew, S.E. Yong, M.P. Toh, and V.J. Lee, Presymptomatic Transmission of SARS-CoV-2 - Singapore, January 23-March 16, 2020. MMWR Morb Mortal Wkly Rep, 2020. 69(14): p. 411–415.

9. Arons, M.M., K.M. Hatfield, S.C. Reddy, A. Kimball, A. James, J.R. Jacobs, J. Taylor, K. Spicer, A.C. Bardossy, L.P. Oakley, et al., Presymptomatic SARS-CoV-2 Infections and Transmission in a Skilled Nursing Facility. N Engl J Med, 2020.

10. Goyal, P., J.J. Choi, L.C. Pinheiro, E.J. Schenck, R. Chen, A. Jabri, M.J. Satlin, T.R. Campion, Jr., M. Nahid, J.B. Ringel, et al., Clinical Characteristics of Covid-19 in New York City. N Engl J Med, 2020.

11. Wang, Z., B. Yang, Q. Li, L. Wen, and R. Zhang, Clinical Features of 69 Cases with Coronavirus Disease 2019 in Wuhan, China. Clin Infect Dis, 2020.

12. Grasselli, G., A. Zangrillo, A. Zanella, M. Antonelli, L. Cabrini, A. Castelli, D. Cereda, Coluccello, G. Foti, R. Fumagalli, et al., Baseline Characteristics and Outcomes of 1591 Patients Infected With SARS-CoV-2 Admitted to ICUs of the Lombardy Region, Italy. JAMA, 2020.

13. Wolfel, R., V.M. Corman, W. Guggemos, M. Seilmaier, S. Zange, M.A. Muller, D. Niemeyer, T.C. Jones, P. Vollmar, C. Rothe, et al., Virological assessment of hospitalized patients with COVID-2019. Nature, 2020.

14. Nguyen, T.H.D. and D.C. Vu, Summary of the COVID-19 outbreak in Vietnam - Lessons and suggestions. Travel Med Infect Dis, 2020: p. 101651.

15. Dinh, L., P. Dinh, P.D.M. Nguyen, D.H.N. Nguyen, and T. Hoang, Vietnam’s response to COVID-19: Prompt and proactive actions. J Travel Med, 2020.

16. Corman, V.M., O. Landt, M. Kaiser, R. Molenkamp, A. Meijer, D.K.W. Chu, T. Bleicker, S. Brünink, J. Schneider, M.L. Schmidt, et al., Detection of 2019 novel coronavirus (2019-nCoV) by real-time RT-PCR. Eurosurveillance, 2020. 25(3).

17. Clinical management of severe acute respiratory infection when COVID-19 is suspected. WHO Interim guidance, 13 March 2020.

18. Team, R.C., R: A language and environment for statistical computing.. R Foundation for Statistical Computing, Vienna, Austria. URL https://www.R-project.org/. 2019.

19. Thanh, H.N., T.N. Van, H.N.T. Thu, B.N. Van, B.D. Thanh, H.P.T. Thu, A.N.T. Kieu, N.N. Viet, G.B. Marks, G.J. Fox, et al., Outbreak investigation for COVID-19 in northern Vietnam. Lancet Infect Dis, 2020.

20. To, K.K., O.T. Tsang, C. Chik-Yan Yip, K.H. Chan, T.C. Wu, J.M.C. Chan, W.S. Leung, T.S. Chik, C.Y. Choi, D.H. Kandamby, et al., Consistent detection of 2019 novel coronavirus in saliva. Clin Infect Dis, 2020.

21. To, K.K.-W., O.T.-Y. Tsang, W.-S. Leung, A.R. Tam, T.-C. Wu, D.C. Lung, C.C.-Y. Yip, J.-P. Cai, J.M.-C. Chan, T.S.-H. Chik, et al., Temporal profiles of viral load in posterior oropharyngeal saliva samples and serum antibody responses during infection by SARS-CoV-2: an observational cohort study. The Lancet Infectious Diseases, 2020.

22. Xu, R., B. Cui, X. Duan, P. Zhang, X. Zhou, and Q. Yuan, Saliva: potential diagnostic value and transmission of 2019-nCoV. Int J Oral Sci, 2020. 12(1): p. 11.

23. Okba, N.M.A., M.A. Muller, W. Li, C. Wang, C.H. GeurtsvanKessel, V.M. Corman, M.M. Lamers, R.S. Sikkema, E. de Bruin, F.D. Chandler, et al., Severe Acute Respiratory Syndrome Coronavirus 2-Specific Antibody Responses in Coronavirus Disease 2019 Patients. Emerg Infect Dis, 2020. 26(7).

24. Inui, S., A. Fujikawa, M. Jitsu, N. Kunishima, S. Watanabe, Y. Suzuki, S. Umeda, and Y. Uwabe, Chest CT Findings in Cases from the Cruise Ship “Diamond Princess” with Coronavirus Disease 2019 (COVID-19). Radiologu: Cardiothoracic Imaging, 2020. In press.

